# GWAS including 82,707 subjects identifies functional coding variant in *OPRM1* gene associated with opioid use disorder

**DOI:** 10.1101/19007039

**Authors:** Hang Zhou, Christopher T. Rentsch, Zhongshan Cheng, Rachel L. Kember, Yaira Z. Nunez, Janet P. Tate, Cecilia Dao, Ke Xu, Renato Polimanti, Lindsay A. Farrer, Amy C. Justice, Henry R. Kranzler, the VA Million Veteran Program, Joel Gelernter

## Abstract

One way to address the current crisis in opioid use is to improve our understanding of the biological mechanisms of opioid use disorder (OUD). We completed a primary GWAS of electronic health record-defined OUD in European-ancestry participants in the Million Veteran Program (MVP) sample, which included 8,529 affected subjects and 71,200 opioid-exposed controls. In the MVP alone, there were no genome-wide significant (GWS) associations. We then subjected the MVP and additional OUD GWAS results from the Yale-Penn and SAGE samples to meta-analysis (in total, 10,544 OUD cases and 72,163 opioid-exposed controls). A functional coding variant (rs1799971, encoding Asn40Asp) in *OPRM1* (mu opioid receptor gene, the main biological target for opioid drugs) reached GWS (p=1.51×10^−8^); then replicated in two independent samples (each at p<0.05). The final meta-analyzed p-value for this variant in all samples was 7.81×10^−10^. SNP-based heritability of OUD was 11.3%. OUD was genetically correlated with 83 traits, including multiple substance use traits, psychiatric illnesses, cognitive performance, and others. Mendelian Randomization revealed possible causal effects on OUD risk from tobacco smoking, major depression, neuroticism, and cognitive performance. Despite the inclusion of data from the MVP, discovery of a significant association depended on including other purpose-collected samples as well. Recruitment of additional opioid dependent subjects for future studies – especially of non-European ancestry – is a crucial next step.

## Introduction

Opioid abuse, addiction, and overdose are at epidemic levels in the United States. Opioids are the leading cause of overdose deaths, and their use has increased dramatically in recent decades [1]. A multifaceted approach is needed to address the opioid crisis, including improving our understanding of the biological mechanisms of opioid addiction. Opioids exert their biological effects primarily by binding (mainly in brain and peripheral nervous tissues) to the opioid receptors *mu* (μ), *kappa* (κ), and *delta* (δ), which are encoded by *OPRM1, OPRK1*, and *OPRD1*, respectively [2]. Numerous candidate-gene association studies of these genes (especially *OPRM1*) and those encoding related proteins have been conducted in the past two decades (reviewed in ref [3, 4]), but prior studies have failed consistently to demonstrate association (e.g., studies on *OPRM1*\*rs1799971 are reviewed in ref [5]). However, a prior genome-wide association study (GWAS) of opioid dosing reported a genome-wide significant (GWS) association mapping upstream of the *OPRM1* locus [6].

Rs1799971 (A118G, encoding Asn40Asp) [5, 7], a functional variant, is one of the most studied candidate variants for substance use traits. While no consistent results were observed, this could reflect limited power and population heterogeneity of previous studies (allele frequency varies greatly in different populations [8, 9]). Several kinds of evidence support possible functional effects of this SNP: rs1799971 reportedly alters beta-endorphin binding and activity [7], may be associated with cortisol response to naloxone blockade [10], and may be associated with neurobehavioral functions in a mouse model [11, 12] and human induced pluripotent stem cell lines [13].

Several GWAS of DSM-IV opioid dependence [OD] yielded significant findings [14-16]; one included internal replication [16] but none reported clear external replication, probably due to the limited sample sizes available (the largest study so far included 2,015 OD cases [16]). The risk variants identified map to *APBB2, PARVA, KCNC1*, and *KCNC2* [14] in African-American (AA) samples, and *CNIH3* [15] and *RGMA* [16] in European ancestry samples. There have also been GWAS of related traits including therapeutic opioid dose (noted above) [6], and opioid overdose (which identified one variant near *MCOLN1* in AAs) [17]. Of these, only the study on opioid dosing included an external validation. No GWAS yet has been sufficiently powered to estimate the SNP-based heritability (*h*^2^) of OD.

We conducted GWAS on ICD (International Classification of Diseases)–9/10-diagnosed opioid use disorder (OUD) and opioid-exposed controls) in 79,729 European Americans (EAs) from the Million Veteran Program (MVP). Then we meta-analyzed for OUD combining MVP, Yale-Penn, and the Study of Addiction: Genetics and Environment (SAGE) samples [18]. The latter two samples were included in our previous publication [16], but were reanalyzed here as a binary diagnostic trait rather than a criterion count for better congruence with available MVP information. Rs1799971 was the only variant that was GWS (p=1.51×10^−8^) in this meta-analysis. We then replicated the result in two independent samples. We estimated the *h*^2^ and detected genetic correlations between OUD and a variety of psychiatric traits. Causal effects of substance use, psychiatric diseases, and educational attainment on liability to OUD were also detected.

## Methods

### MVP datasets

The MVP is a cross-sectional mega-biobank supported by the U.S. Department of Veterans Affairs (VA). Enrollment in MVP began in 2011 and is ongoing. Phenotypic data were collected using the VA electronic health record (EHR), and blood samples were obtained for genetic studies [19]. Two phases of genotypic data have been released according to their genotyping epochs and were included in this study. MVP phase1 contains 353,948 subjects, of whom 209,020 were defined previously as EAs [20]. MVP phase2 contains 108,416 subjects. We used the same process as in MVP phase1 for quality control and to define EAs (see Supplementary Methods) [20]; there were 67,268 EA subjects.

Cases were participants with at least one inpatient or two outpatient ICD-9/10 codes for OUD (Supplementary Table 1) between 2000 and 2018. In MVP phase1, there were 6,367 OUD EAs (3.04% prevalence), and 2,162 OUD EAs (3.21% prevalence) in MVP phase2 were included in this study. Stringent criteria were applied to define incident opioid-exposed controls (see details in ref [21]). In short, we started with all MVP participants and excluded subjects with exposure to a prescription opioid <7 consecutive days, or with VA follow-up less than 6 months after baseline, or with cancer diagnosed before or after baseline, or with baseline opioid dosage >90 mg morphine equivalent daily dose (MEDD), or with OUD diagnosis or OUD treatment at baseline. For the remaining participants, a latent growth mixture model was applied to identify the major classes of opioid dose (measured by MEDD) trajectories that assigned each individual to the trajectory with the highest probability of membership. Four resultant MEDD trajectories were designated as low, moderate, escalating, and rapidly escalating. To minimize the potential rate of false negatives in the control group, subjects assigned to the low-dose trajectory without an incident OUD diagnosis during follow-up were defined as controls, yielding 55,429 and 15,771 EA controls in MVP phaes1 and MVP phase2, respectively.

Genotyping in MVP was performed using a customized Affymetrix Biobank Array. Imputation and quality control metrics for MVP phase1 were as described previously [20]. Similar processes were used for MVP phase2 (see Supplementary Methods). GWAS was then performed on the MVP datasets. We used logistic regression implemented in PLINK v1.90b4.4 [22] for the OUD GWAS correcting for age, sex, and the first 10 principal components (PCs).

### Ethics statement

The Central VA Institutional Review Board (IRB) and site-specific IRBs approved the MVP study. All relevant ethical regulations for work with human subjects were followed in the conduct of the study, and written informed consent was obtained from all participants.

### Yale-Penn and SAGE datasets

GWAS for DSM-IV OD criterion counts were performed previously, including three phases of Yale-Penn data, and the SAGE cohort (dbGaP study id phs000092.v1.p1) [16]. We re-analyzed these data using OUD diagnosis. See Supplementary Methods.

### Meta-analyses

Sample-size-weighted meta-analyses were performed using METAL [23]. Given the unbalanced ratios of cases to controls in MVP samples, effective sample sizes were calculated as follow:

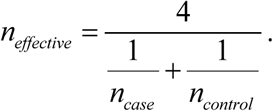

The calculated effective sample sizes in MVP were used in meta-analyses and all downstream analyses. Only variants present at least in MVP phase1, which is the largest sample (∼75% of the total), and with heterogeneity test p-value >5×10^−8^ were retained, leaving 5.7 M variants.

### Replication in independent samples

In Yale-Penn, 4,817 subjects were recently added and not included in any prior analysis. We genotyped them using the Illumina Multi-Ethnic Genotyping Array (San Diego, CA) which includes ∼1.7 M SNPs. Subjects with mismatched genotypic and phenotypic sex were removed, as were subjects with excessive heterozygosity. Duplicate subjects with respect to the Yale-Penn discovery samples were removed. The remaining subjects were classified into population groups as for MVP. Among the 2,041 genetically classified EAs, 508 were diagnosed as DSM-IV OD cases, and 206 were opioid exposed controls. GEMMA was used for an association test only for rs1799971 (i.e., no other markers were evaluated) and corrected for age, sex, and the first 10 PCs.

In the UK Biobank (UKB), we looked up the association between rs1799971 (only this marker, as for the other replication sample) and buprenorphine treatment (mostly used to treat OUD; treatment/medication code: 20003_1140871732) in the UKB. We examined GWAS summary data released by the Neale lab (information available at http://www.nealelab.is/uk-biobank) for 240 cases and 360,901 controls differentiated based on buprenorphine treatment.

### SNP-based *h*^2^

LD Score Regression (LDSC) [24] was used to estimate the SNP-based *h*^2^ using 1000 Genomes Project Europeans [8] as the LD reference panel. The major histocompatibility complex (MHC) region (chr6: 26–34Mb) was excluded. Effective sample size was used in LDSC.

### Genetic correlation

We estimated the genetic correlation (*r*_g_) between OUD and 715 publicly available traits from LD Hub [25] or other resources using LDSC (Supplementary Table 2) [26]. Among the tested traits, 232 were published previously, and 483 from the UKB were unpublished, but integrated in LD Hub. Bonferroni correction was applied and correlation was considered significant at a p-value threshold of 6.99×10^−5^.

### Mendelian Randomization

We used Mendelian randomization (MR) to investigate whether exposures (based on 18 published traits that were significantly correlated with OUD [*r*_g_ p<6.99×10^−5^]) have causal effects on the liability to OUD. For instrumental variants missing in the OUD summary data, we used the results of the best proxy variant in highest LD (*r*^2^>0.8) with the missing variant. If the MAF of the missing variant was <0.01, or none of the variants within 200 kb had LD *r*^2^>0.8, we removed the instrumental variant from the analysis. Palindromic SNPs (A/T or G/C alleles) with MAF [0.4, 0.5] in the OUD summary data were also removed or replaced with the best proxy variant. For robust causal effect inference, we limited the traits studied to those with >30 available instruments. Accordingly, 12 exposures were analyzed. We used weighted median [27], inverse-variance weighted (IVW, random-effects model) [28], and MR-Egger [29], implemented in the R package “MendelianRandomization v0.3.0” [30] for MR inference. Evidence of pleiotropic effects was examined by the MR-Egger intercept test, where a non-zero intercept (p<0.05) indicates directional pleiotropy [29]. Whereas MR analyses require the beta (effect size) and standard error, we calculated these using Z-scores (*z*), allele frequency (*p*) and sample size (*n*) from the OUD meta-analyses [31]:

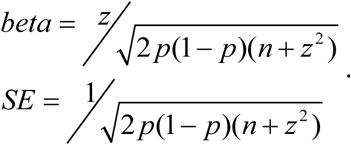

## Results

### Association results for opioid use disorder (OUD)

In MVP phase1, 6,367 subjects were diagnosed as OUD cases, and 55,429 subjects were defined as controls. 2,162 cases and 15,771 controls were included from MVP phase2 (Table 1). We meta-analyzed the 8,529 OUD cases and 71,200 controls within MVP (totaling 79,729 individuals, Table 1), and no variant reached GWS (p<5×10^−8^, Supplementary Figure 1). The variant with the smallest p-value was rs1799971 in the *OPRM1* gene (p=5.90×10^−8^, *n*_effective_=30,443; the minor G allele is protective with beta=-0.142 and se=0.026).

**Table 1.**
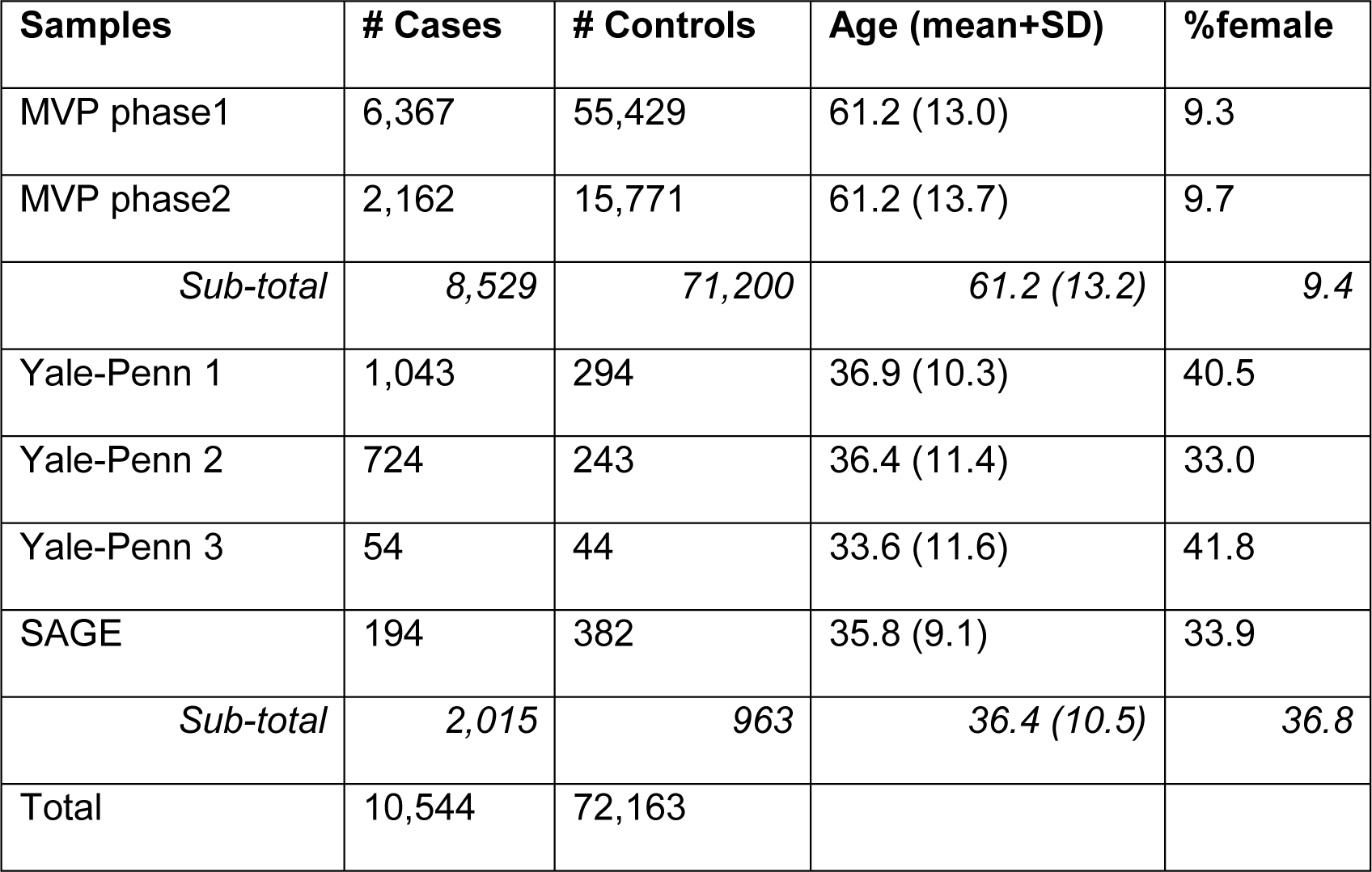
Demographics – discovery sample.

We then meta-analyzed the MVP samples with Yale-Penn (three tranches) and SAGE samples, bringing the total sample size to 82,707 (10,544 cases and 72,163 opioid-exposed controls, Table 1). This represents a 24% increase in the number of cases. The OD cases in Yale-Penn and SAGE were diagnosed using DSM-IV, and the controls were opioid exposed (this differs from our previously published GWAS [16], which defined phenotype based on DSM-IV OD criterion count). From the meta-analysis, SNP-based heritability (*h*^2^) was 0.113 (se=0.018) estimated by LD Score Regression (LDSC). The association of rs1799971 with OUD was GWS (beta=-0.066, se=0.012, p=1.51×10^−8^, *n*_effective_=33,421, Figure 1, Supplementary Figure 2). The effects were all in the same direction except for SAGE, which might be due to limited sample size. There were no significant results from gene-based association and gene-set analyses.

**Figure 1.**
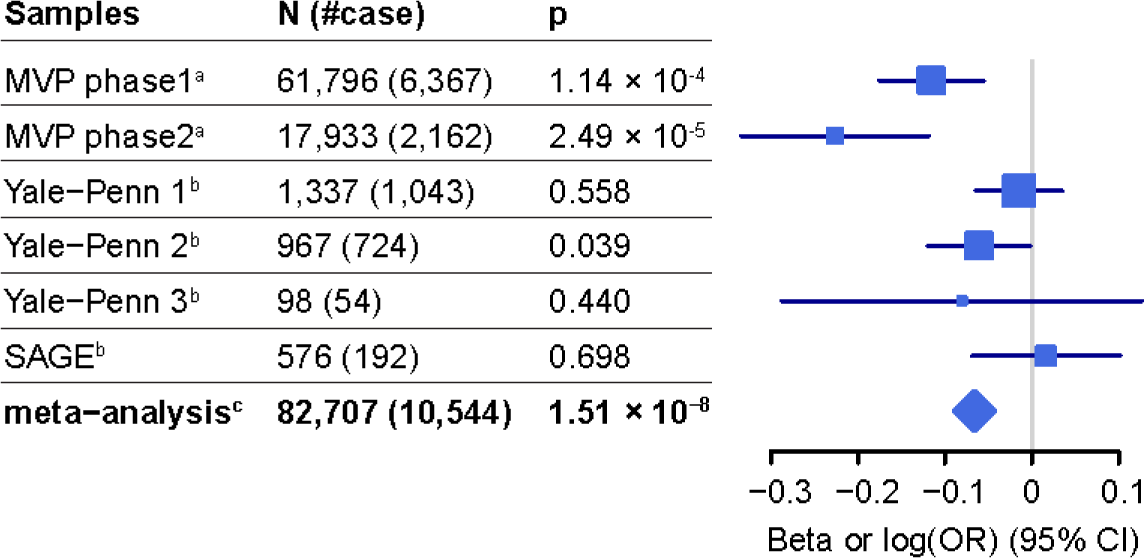
Associations between rs1799971*G and OUD. ^a^Logistic regression was applied on unrelated case/control samples in MVP, log(OR) is presented; ^b^a linear mixed model was applied on complex family-based samples, beta is presented; ^c^effective sample size weighted meta-analysis was applied, and beta is presented.

**Figure 2.**
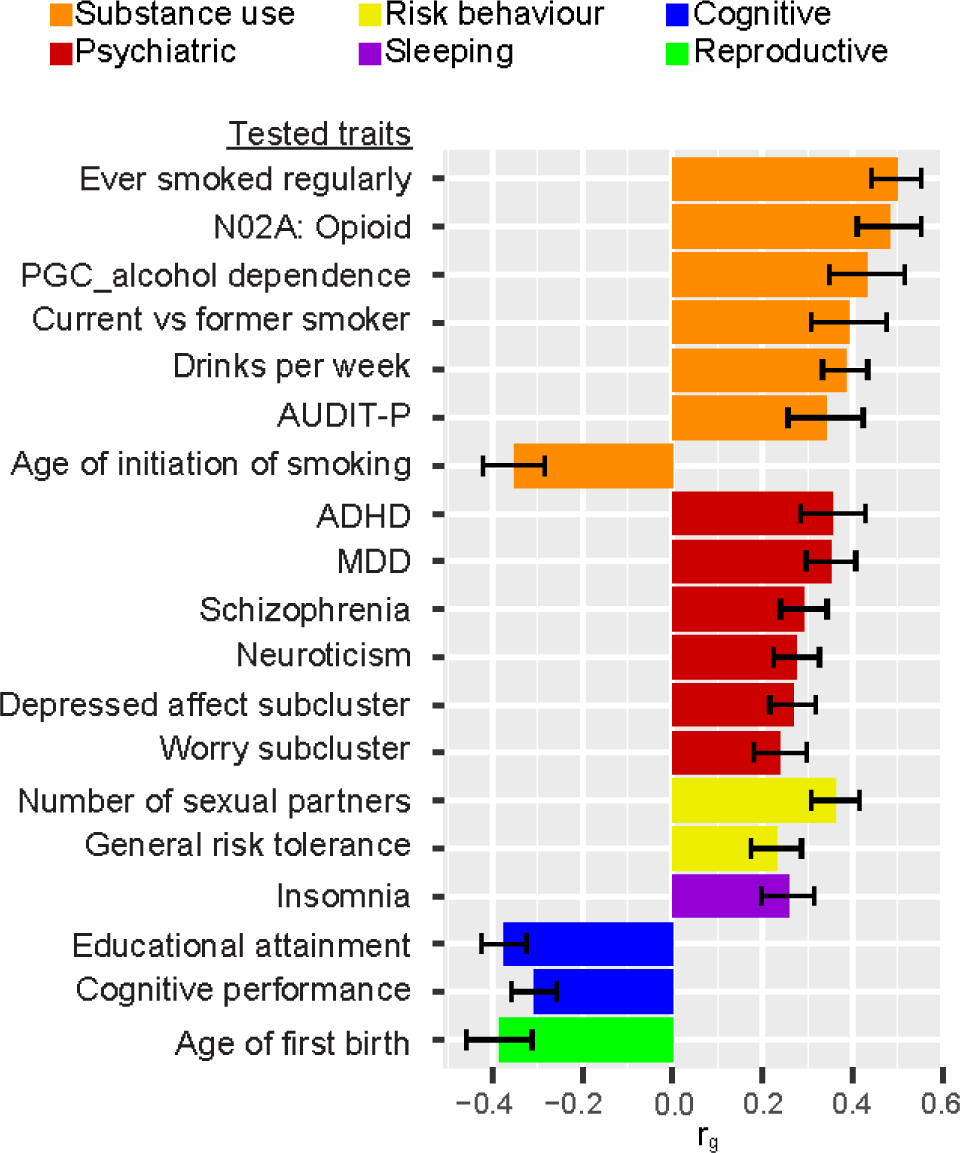
Genetic correlations between OUD and published traits. Listed are the 18 published traits significantly correlated with OUD. *N02A: Opioid*: self-reported medication-use of opioid drugs (Anatomical Therapeutic Chemical [ATC] Classification code: N02A) in UK Biobank; *PGC*: Psychiatric Genomics Consortium; *AUDIT-P*: the Alcohol Use Disorders Identification Test–Problems; *ADHD*: attention deficit hyperactivity disorder, *MDD*: major depressive disorder; *Depressed affect subcluster*: depressed affect neuroticism subcluster; *Worry subcluster*: worry neuroticism subcluster.

### Replication in independent samples

In total, 714 EAs (508 OD cases and 206 opioid-exposed controls) were analyzed in the new Yale-Penn samples, and rs1799971*G was associated with reduced OD risk (i.e., in the same direction as the discovery meta-analysis) (beta=-0.074, se=-0.038, p=0.049). In the UKB, rs1799971*G was negatively associated with buprenorphine treatment status (240 cases and 360,901 controls, beta=-1.90×10^−4^, se=9.13×10^−5^, p=0.038), also consistent with the direction of effect in the discovery sample. Meta-analysis of discovery and replication cohorts for this variant yielded a p-value of 7.81×10^−10^.

### Genetic correlations with other traits

We estimated the genetic correlations (*r*_g_) between OUD and 715 traits with publicly available summary statistics using LDSC. Among those traits, 232 were published, and 483 were unpublished and derived from UKB data. OUD was significantly correlated with 83 of these traits (Supplementary Table 2). Figure 1 depicts 18 correlated traits from published literature (Supplementary Methods). Among the correlated substance use-related traits, “ever smoked regularly” showed the highest correlation with OUD (*r*_g_=0.51, se=0.06, p=3.37×10^−19^), followed by “opioid medication use” in UKB (*r*_g_=0.48, se=0.07, p=1.61×10^−11^). Both “problematic alcohol use” (measured by alcohol dependence and AUDIT-P [Alcohol Use Disorders Identification Test–Problems] score) and “alcohol use quantity” (measured by drinks per week) showed high genetic correlations with OUD. “Unable to stop smoking” (current *vs*. former smoker), and “earlier age of smoking initiation” were also correlated with OUD. However, correlations with AUDIT-C (Alcohol Use Disorders Identification Test–Consumption), total AUDIT, cigarettes per day, and lifetime cannabis use were not significant after Bonferroni correction. Several psychiatric traits were correlated with OUD, including attention deficit hyperactivity disorder (ADHD, *r*_g_=0.36, se=0.07, p=6.78×10^−7^), major depressive disorder (MDD, *r*_g_=0.35, se=0.06, p=1.62×10^−10^), schizophrenia (*r*_g_=0.29, se=0.05, p=1.93×10^−8^), neuroticism (*r*_g_=0.27, se=0.05, p=8.65×10^−8^), and neuroticism subclusters. OUD was positively correlated with risk-taking behavior and insomnia, and negatively correlated with cognitive traits and age of first birth. These finding are consistent with the known adverse medical, psychiatric, and social consequences of OUD.

### Mendelian Randomization

Using MR, we explored possible causal effects of exposures on OUD (Table 2). Among the 12 tested exposures, five supported a possible causal effect on liability to OUD by at least one method and without evidence of horizontal pleiotropy (MR-Egger intercept p>0.05): positively with ever smoked regularly, MDD, neuroticism and worry neuroticism subcluster, and negatively with educational attainment. There was weak evidence of a causal effect of drinks per week on OUD risk by the IVW method, but the estimate could be biased due to horizontal pleiotropy.

**Table 2.** Causal effects on OUD by MR.

| Exposure (#instruments) | IVW |  | Weighted median |  | MR-Egger |  | MR-Egger intercept p |
| --- | --- | --- | --- | --- | --- | --- | --- |
| | $\beta$ (se) | p | $\beta$ (se) | p | $\beta$ (se) | p | |
| **Ever smoked regularly (197)** | 0.33 (0.04) | **$1.88 \times 10^{-19}$** | 0.28 (0.05) | **$4.79 \times 10^{-8}$** | 0.18 (0.14) | 0.206 | 0.264 |
| Drinks per week (63) | 0.31 (0.10) | **$1.31 \times 10^{-3}$** | 0.23 (0.13) | 0.078 | -0.04 (0.16) | 0.820 | $8.77 \times 10^{-3}$ |
| **MDD (78)** | 0.18 (0.05) | **$6.21 \times 10^{-4}$** | 0.17 (0.06) | $4.28 \times 10^{-3}$ | -0.25 (0.34) | 0.468 | 0.202 |
| Schizophrenia (108) | 0.03 (0.01) | 0.020 | 0.03 (0.02) | 0.097 | -0.01 (0.07) | 0.825 | 0.439 |
| **Neuroticism (131)** | 0.22 (0.06) | **$4.07 \times 10^{-4}$** | 0.16 (0.08) | 0.050 | 0.19 (0.27) | 0.468 | 0.934 |
| Depressed affect subcluster (55) | 0.32 (0.10) | $2.12 \times 10^{-3}$ | 0.34 (0.12) | $4.01 \times 10^{-3}$ | -0.35 (0.50) | 0.482 | 0.173 |
| **Worry subcluster (61)** | 0.28 (0.09) | **$1.32 \times 10^{-3}$** | 0.15 (0.11) | 0.157 | 0.24 (0.40) | 0.545 | 0.919 |
| Number of sexual partners (64) | 0.27 (0.09) | $1.77 \times 10^{-3}$ | 0.18 (0.10) | 0.083 | 0.43 (0.40) | 0.283 | 0.689 |
| General risk tolerance (64) | 0.16 (0.13) | 0.212 | 0.18 (0.16) | 0.257 | 0.45 (0.58) | 0.438 | 0.608 |
| Insomnia (158) | 0.06 (0.02) | $3.45 \times 10^{-3}$ | 0.03 (0.03) | 0.226 | -0.03 (0.09) | 0.731 | 0.275 |
| **Educational attainment (564)** | -0.32 (0.04) | **$2.34 \times 10^{-16}$** | -0.33 (0.05) | **$4.09 \times 10^{-10}$** | -0.18 (0.15) | 0.222 | 0.311 |
| Cognitive performance (133) | -0.12 (0.05) | $9.72 \times 10^{-3}$ | -0.11 (0.06) | 0.061 | 0.03 (0.22) | 0.894 | 0.477 |

P-values in bold are significant after multiple testing correction (significant threshold, $0.05/36=1.39 \times 10^{-3}$ ). Traits labeled in bold are those having a causal effect on OUD by at least one method without evidencing horizontal pleiotropy (MR-Egger intercept $p>0.05$ ). IVW: inverse-variance weighted (IVW) linear regression. MDD: major depressive disorder. Depressed affect subcluster: depressed affect subcluster of neuroticism. Worry subcluster: worry subcluster of neuroticism.

## Discussion

Opioid use is at epidemic levels in the United States and is a major cause of death and disability worldwide. Understanding the genetic architecture of OUD might provide clinically useful clues about its biology. However, only a few risk variants have been identified by GWAS so far, and none has had clear external replication. Several factors contribute to this situation: 1). OUD is a complex psychiatric disease with relatively low heritability, and there is no single variant with large effect size that can be detected in small cohorts (contrary to, for example, alcohol dependence [32] with *ADH1B*, and nicotine dependence with the chromosome 15 nicotine receptor cluster [33]); 2). Previous OUD GWAS were relatively small compared to those for legal substance use disorders (e.g., the number of alcohol use disorder cases reached 57,564 in a large meta-analysis [34]); 3). In published work relevant to opioid use, there was considerable phenotypic heterogeneity across samples. The ascertainment of OUD cases (e.g., ICD-diagnosed OUD in the EHR, DSM-IV-assessed OD, patients receiving opioid substitution therapy, and daily injectors of illicit opioids) and controls (e.g., opioid exposed, or random population with unknown opioid exposure status) differ by study. One way to reach a better understanding of OUD genetics is increasing the sample size in a homogeneous cohort.

We conducted GWAS of OUD in a large cohort, the MVP, comprising 8,529 cases and 71,200 opioid-exposed controls of European ancestry. Most previously reported variants associated with a wide range of opioid-related traits were not significant in MVP (cf. summary statistics). For some, this reflects lack of marker information or LD proxies in the MVP, or the lack of non-European populations in this analysis; some associations were previously reported in African-ancestry populations only [14]; others were reported in EAs, but relevant variants are missing in the MVP data (e.g., rs12442183 near *RGMA* reported by Cheng et al. [16] was filtered by low genotype call rate in imputation). No variant reached GWS in this largest-ever cohort individually; *OPRM1*\*rs1799971 was nominally significantly associated with OUD (p=5.90×10^−8^). We meta-analyzed MVP samples with Yale-Penn and SAGE (re-analyzed here as case/control rather than the quantitative criterion count traits reported originally, to match the available phenotype from the MVP more closely), increasing total sample size to 82,707 (10,544 cases and 72,163 opioid-exposed controls). By adding four samples from Yale-Penn and SAGE, rs1799971 reached GWS. The final meta-analyzed p-value for this marker is 1.51×10^−8^ (excluding independent replications).

Rs1799971 (A118G) maps to exon 1 of the mu opioid receptor (*OPRM1*) gene, causing an amino acid change from asparagine to aspartic acid. Extensive candidate studies of this variant with a wide range of addictive and other behavioral traits have been conducted over two decades. Associations between rs1799971 and opioid-related traits have not been consistent [5]. We conducted hypothesis-free, genome-wide analyses for OUD and detected association at rs1799971 by almost quintupling the number of cases compared to any previous studies [5, 16]. Our increment in exposed controls, which have often been even more limiting than affected OUD subjects in previous studies, is even greater. Since many individuals exposed to opioids become dependent, an unassessed control group is not an ideal alternative to an opioid-exposed control group even if greater numbers of subjects can be achieved – because the former group is more correctly “diagnosis unknown” and inevitably contains many subjects genetically predisposed to OUD who would express that phenotype had they been exposed. We sought replication in two independent samples. One included newly genotyped Yale-Penn subjects, and the other, a proxy-phenotype buprenorphine treatment sample from the UKB. The association was replicated in each of these samples.

Although we still encounter a relative lack of power overall as indicated by no additional variant detected beyond rs1799971, a lack of any significant gene associated in gene-based analysis, and no gene-set enrichment, there is reasonable power for genetic correlation. Multiple substance use-related traits including smoking, alcohol, and opioid use and psychiatric traits were among the top correlates. Several smoking traits were positively correlated with OUD, consistent with the strong correlation between nicotine use and opioid use disorder [35, 36]. MR analysis provided evidence (considered weak, as it is not supported by all the three tested methods) that the genetic liability to substance use related traits has causal effects on susceptibility to OUD. Medical opioid use is correlated with OUD, as expected. And problematic alcohol use (measured by AD and AUDIT-P score) and drinking quantity are also genetically correlated with OUD. Thus, it may be feasible for prevention or treatment efforts directed at legal substance use to reduce the burden of consequent opioid dependence. Psychiatric traits including ADHD, MDD, schizophrenia, and neuroticism are genetically correlated with OUD, consistent with phenotypic evidence [37, 38]. Weak evidence from MR analyses also indicated possible causal effects on OUD risk of MDD and neuroticism.

We note limitations of this study. First, the sample size, though a major improvement, is still not as large as what can be obtained for legal substance use-related traits, and this limited power to detect more GWS signals and to obtain insight into OUD biological mechanisms. Legal substance use traits are more common, and data pertinent to these traits is collected more commonly than for illegal traits in biobanks and EHRs. Second, the phenotypes in the samples we studied were not identical. The MVP used ICD 9/10-diagnosed OUD. We expect false negatives in a sample like the MVP, owing to stigma and insufficient vigilance on the part of treatment teams concentrating mostly on medical illness, but few false positives. Overall, we believe these diagnoses to be quite useful, as false positives would be more of a problem than false negatives. Third, the only GWS variant, rs1799971, has a very small effect size (beta=-0.066). The association was close to GWS in MVP alone, and improved by meta-analysis. High quality genotype data and replication in an independent sample were required. Rs1799971 was genotyped directly (was not imputed) in all samples, discovery and replication. Fourth, the replication samples are small (508 OD cases in the new Yale-Penn sample and 240 buprenorphine treatment cases in UKB), and the associations only nominally significant (p∼0.05). We examined only this one marker in the replication samples, so nominal significance should be a sufficient test of replication (and arguably a 1-tailed test could have been applied rather than the more stringent 2-tailed test applied here). The phenotype in UKB is a proxy phenotype–buprenorphine treatment. Although buprenorphine is a first-line drug for OUD treatment, it could have been used for other purposes in the UKB population, including pain management; but if this is true to any considerable extent, it should reduce our power to detect an association due to added noise, rather than lead to a false positive finding. Fifth, we only studied samples with European ancestry. There has been a lack of recruitment for non-European populations globally, e.g., only a few GWASs have been conducted in AAs [6, 14] in smaller cohorts.

In summary, we report here the largest GWAS and the largest meta-analysis for OUD so far. This finding may not have direct implications for personalized medicine – because the relevant gene is already the main physiological target of all opioids, illegal and therapeutic; providing, at least, a “proof of principle” of relevance of the finding. OUD is genetically correlated with substance use traits, other psychiatric traits, insomnia, and cognitive performance. Among these, ever smoked regularly, MDD, neuroticism, and cognitive performance have potential causal or protective effects on the liability of OUD, which provides clues for future prevention efforts. Recruitment of additional OUD subjects – especially of non-European ancestry – is a crucial next step. Considering the general lack of private foundation funding for study of substance use disorders, it is likely that government-supported funding agencies will be required to accomplish this goal.

## Data Availability

The summary statistics will be available after publication.

## Acknowledgements

This research used data from the Million Veteran Program, Office of Research and Development, Veterans Health Administration, and was supported by award #1I01BX003341. This publication does not represent the views of the Department of Veterans Affairs or the United States Government. Supported also by a NARSAD Young Investigator Grant from the Brain & Behavior Research Foundation (HZ). Dr. Gelernter was supported by National Institutes of Health grants RC2 DA028909, R01 DA12690, R01 DA12849, R01 DA18432, R01 AA11330, and R01 AA017535, the New England MIRECC, and by the Department of Veterans Affairs Medical Research Program. Dr. Kranzler was supported by National Institutes of Health grants R21 DA10242 and R01 DA18432 and both he and Dr. Kember were supported by the VISN 4 MIRECC. Genotyping services for a part of the Yale-Penn GWAS study were provided by the Center for Inherited Disease Research (CIDR) and Yale University (Center for Genome Analysis). CIDR is fully funded through a federal contract from the National Institutes of Health to The Johns Hopkins University (contract number N01-HG-65403).

The publicly available dataset of Study of Addiction: Genetics and Environment (SAGE) used for the analysis was obtained from dbGaP at https://www.ncbi.nlm.nih.gov/projects/gap/cgi-bin/study.cgi?study_id=phs000092.v1.p1. Funding support for SAGE was provided through the NIH (U01 HG004422). SAGE is one of the genome-wide association studies funded as part of the Gene Environment Association Studies (GENEVA) under Genes, Environment and Health Initiative (GEI). Assistance with phenotype harmonization and genotype cleaning, as well as with general study coordination, was provided by the GENEVA Coordinating Center (U01 HG004446).

We also thank Ann Marie Lacobelle, and Christa Robinson who provided technical assistance for the genotyping.

